# Modelling, Simulations and Analysis of the First and Second COVID-19 Epidemics in Beijing

**DOI:** 10.1101/2021.07.04.21259205

**Authors:** Lequan Min

## Abstract

**Background:** In December 2019, a novel coronavirus-induced pneumonia (COVID-19) broke out in Wuhan, China. On 19 January and on 8 June 2020, there were two wave COVID-19 epidemics happened in Beijing. Modelling, simulations and analysis for the two wave epidemics are important issues.

**Methods:** This study introduces a symptomatic-asymptomatic-recoverer-death differential equation model (SARDDE). It presents the conditions of the asymptotical stability on the disease-free equilibrium of the SARDDE. It proposes the necessary conditions of disease spreading for the SARDDE. Based on the reported data of the first and the second COVID-19 epidemics in Beijing and numerical simulations, it determines the parameters of the SARDDE, respectively.

**Results:** Numerical simulations of the SARDDE describe well the outcomes of the current symptomatic and asymptomatic individuals, the recovered symptomatic and asymptomatic individuals, and the died infected individuals, respectively. The numerical simulations obtain the following results.

- The transmission rate of the symptomatic infections caused by the symptomatic individuals in the second Beijing epidemic is about two times higher than the one in the first Beijing epidemic.
- Both the symptomatic and the asymptomatic individuals cause lesser asymptomatic spread than symptomatic spread.
- The blocking rates of 89.32% and 97.48% (reaching the infection turning points) to the symptomatic infections cannot prevent the spreads of the first and the second COVID-19 epidemics in Beijing, respectively.
- That on the day 28, the symptomatic infection blocking rates reached to 100% has made the second Beijing epidemics epidemic end on day 56.
- That on the day 98, the symptomatic infection blocking rates reached to 100% has made the the first Beijing epidemics epidemic end on day 140.
- Keeping the blocking rates, the recovery rates and the death rates reaching the infection turning points would make the numbers of current hospitalized infected individuals reach, on day 140, 208958 individuals and 25017 individuals for the two Beijing epidemics, respectively.

**Conclusions:** Virtual simulations suggest that the strict prevention and control strategies implemented by Beijing government are effective and necessary; using the data from the beginning to the days after about two weeks after the turning points can estimate well and approximately the following outcomes of the two COVID-19 epidemics, respectively. To avoid multiple epidemic outbreaks, a recommendation is that the authorities need to have maintained the prevention and control measures implemented, at least, 7 days after reaching the turning point until new current infection cases disappear. It is expected that the research can provide better understanding, explaining, and dominating for epidemic spreads, prevention and control measures.

## 1 Introduction

In December 2019, a novel coronavirus-induced pneumonia (COVID-19) broke out in Wuhan, China. As of 15 July 2022, over 557 million confirmed cases of COVID-19 and over 6.35 million deaths have been reported globally [1]. Now COVID-19 affects more than 220 countries and regions including Antarctica.

One of the reasons of such a tragedy is that people in some countries do not pay attentions to theoretical analysis and estimations for COVID-19 epidemics. In fact mathematical models for epidemic infectious diseases have played important roles in the formulation, evaluation, and prevention of control strategies. Modelling the dynamics of spread of disease can help people to understand the mechanism of epidemic diseases, formulate and evaluate prevention and control strategies, and predict tools for the spread or disappearance of an epidemic [2].

Since the outbreak of COVID-19 in Wuhan, many scholars have published a large numbers of articles on the modeling and prediction of COVID-19 epidemic (for examples see [3–15]). It is difficult to describe well the dynamics of COVID-19 epidemics. In a Lloyd-Smith et al’s paper, it described nine challenges in modelling the emergence of novel pathogens, emphasizing the interface between models and data [16].

On 19 January, two Beijingers returning from Wuhan were diagnosed with COVID-19. That triggered the first wave of COVID-19 in Beijing. During the first wave of COVID-19, a total of 420 locally diagnosed cases were reported. After 140 days, on 8 June, 411 COVID-19 infected individuals were cured and 9 died [17]. However three days later, there was one new confirmed individual infected a kind of different COVID-19 in an agricultural products wholesale market in Tongzhou district, Beijing. This triggered a second wave of COVID-19 epidemic in Beijing. After 56 days, totals of 335 and 50 locally symptomatic and asymptomatic COVID-19 individuals were reported, and all individuals were cured. Medical staff have achieved zero infection [17].

This paper introduces a SARDDE. It gives the conditions of the asymptotical stability on the disease-free equilibrium of the SARDDE. Using simulations determines the parameters of the SARDDE based on the reported data of the two COVID-19 epidemics in Beijing [17]. Numerical simulations of the SARDDE describe well the practical outcomes of current infected symptomatic and asymptomatic individuals, recovered infected symptomatic and asymptomatic individuals, and died infected individuals. Virtual simulations are given to estimate the effectiveness of the prevention and control strategies. This paper improves the simulation results implemented in the previous version of this paper [18], and increases a comparing subsection.

The rest of this article is organized as follows. Subsections 2.2.1 and 2.3.1 establish the SARDDEs with 5 and 4 variables, respectively. Subsections 2.2.2 and 2.3.2 provide the criterions of the asymptotical stability of the disease-free equilibriums of the two SARDDEs. Subsections 2.2.3 and 2.3.3 determine the necessary conditions of disease spreading. Subsections 3.1.1 and 3.2.1 implement the dynamic simulations of the SARDDEs to describe the reported data of the first and second COVID-19 epidemic in Beijing, respectively. Subsections 3.1.2 and 3.2.2 discuss and analyze the simulation results. Subsections 3.1.3 and 3.2.3 implement virtual simulation examples to predict the outcomes of the two Beijing epidemics on different prevention and control measures. Subsection 3.3 compares the two COVID-19 epidemics in Beijing. Section 4 concludes the paper. A recommendation to avoid multiple epidemic outbreaks is proposed in the same Section.

## 2 SARDDEs and Dynamic Properties

### 2.1 Data collection

The original COVID-19 data sets of the first and second epidemics in Beijing were downloaded from the Beijing authority’s Website [17]. The author edited the original COVID-19 data sets for using in this study.

### 2.2 SARDDE with death variable

#### 2.2.1 Model

For SARDDE model with death variable, there are five states. *I*(*t*), *I*_*a*_(*t*), *I*_*r*_(*t*), *I*_*ra*_(*t*) and *D*(*t*) represent the fraction of current symptomatic infected individuals, and current asymptomatic infected individuals, cumulative recovered symptomatic infected individuals, cumulative recovered asymptomatic infected individuals and cumulative died individuals, respectively. The transition among these states is governed by the following rules (Flowchart of the rules is shown in Fig.1, where *S* represents susceptible population.).

**Figure 1.**
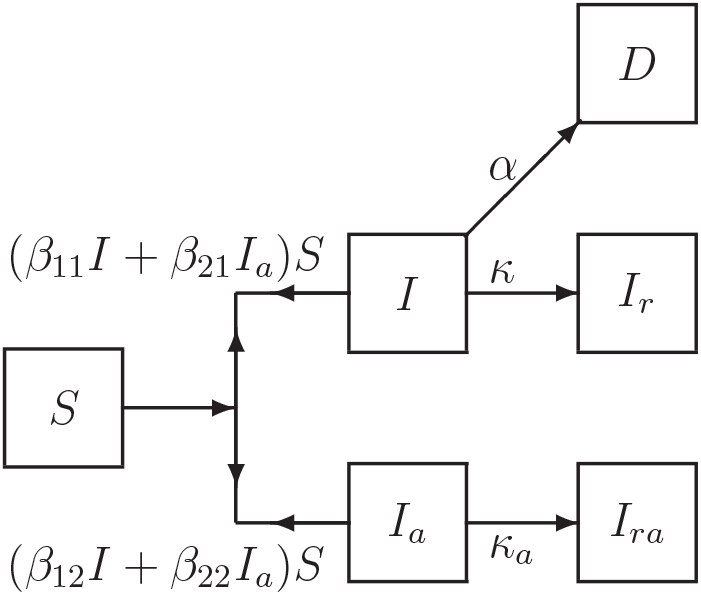
Flowchart of disease transmission among susceptible population *S*, current symptomatic infected individuals *I*, current infected individuals *I*_*a*_, recovered symptomatic infected individuals *I*_*r*_, recovered asymptomatic but infected individuals *I*_*ra*_, and died individuals *D*.

First, the symptomatic infected individuals (*I*) and the asymptomatic infected individuals (*I*_*a*_) infect the susceptible population (*S*) with the transmission rates of *β*_11_ and *β*_21_, respectively, making *S* become symptomatic infected individuals, and with the transmission rates of *β*_12_ and *β*_22_, respectively, making *S* become asymptomatic individuals. Then, a symptomatic individual is cured at a rate *κ*, an asymptomatic individual returns to normal at a rate *κ*_*a*_. An infected individual dies at a rate *α*. Here all parameters are positive numbers.

Assume that the dynamics of an epidemic can be described by *m*-time intervals, which correspond different prevention and control measures, and medical effects. At *l*th time interval, the SARDDE has the form:

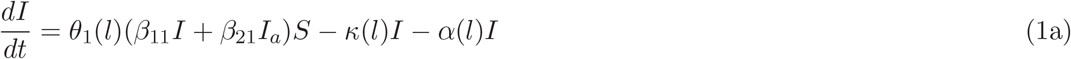

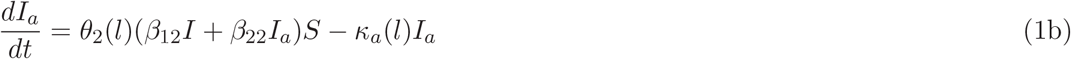

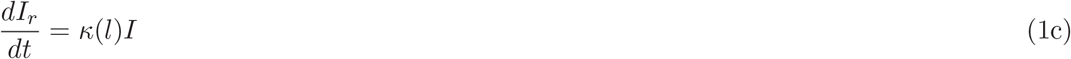

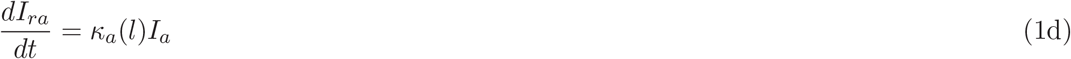

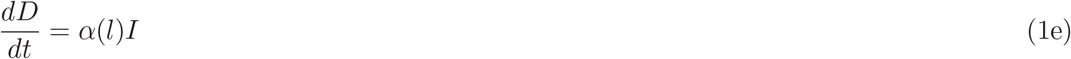

where Θ_1_(*l*) = (1 − *θ*_1_(*l*)) and Θ_2_(*l*) = (1 − *θ*_2_(*l*)) (*l* = 1, …, *m*) represent the blocking rates to symptomatic and asymptomatic infections, respectively. Then SARDDE (1) has a disease-free equilibrium:

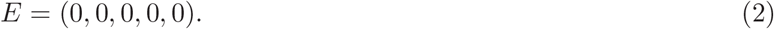

#### 2.2.2 Stability of disease-free equilibrium

The stability of SARDDE (1) is determined by the first two equations (1a) and (1b). Denote in (1a) and (1b)

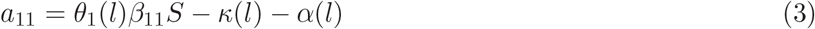

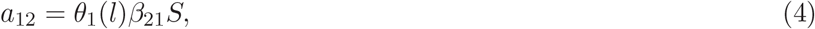

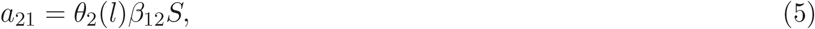

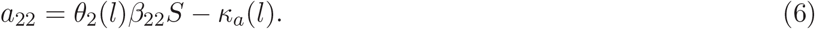

Then at the disease-free equilibrium of SARDDE (1), the Jacobian matrix of (1a) and (1b) is

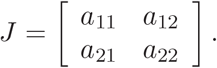

Solving the corresponding eigenequation obtains two eigenvalues:

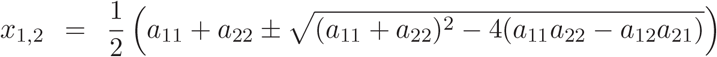

Therefore we obtain the following:

##### Theorem 1

*Suppose that a*_11_, *a*_12_, *a*_21_ *and a*_22_ *are defined by (3)-(6). Then the disease-free equilibrium E (2) of SARDDE (1) is globally asymptotically stable if, and only if, the following inequalities hold:*

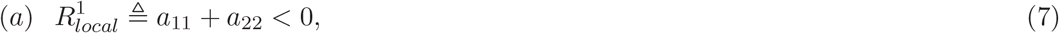

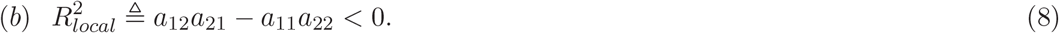

#### 2.2.3 The necessary condition of disease spreading

If an epidemic can occur, then

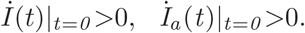

This implies that

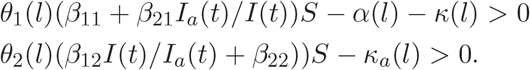

Solving the above inequalities gives the following

##### Theorem 2

*If SARDDE (1) satisfies the following inequalities*

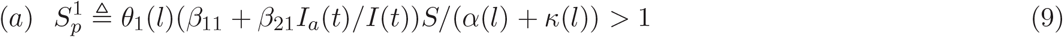

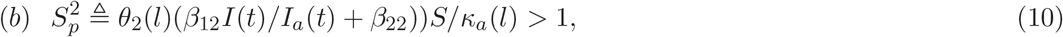

*then a disease transmission will occur*.

### 2.3 SARDDE Model without death variable

#### 2.3.1 Model

Similar to Section 2.2.1, the transition among these states is governed by the following rules (Flowchart of the rules is shown in Fig. 2, where *S* represents susceptible population.)

**Figure 2.**
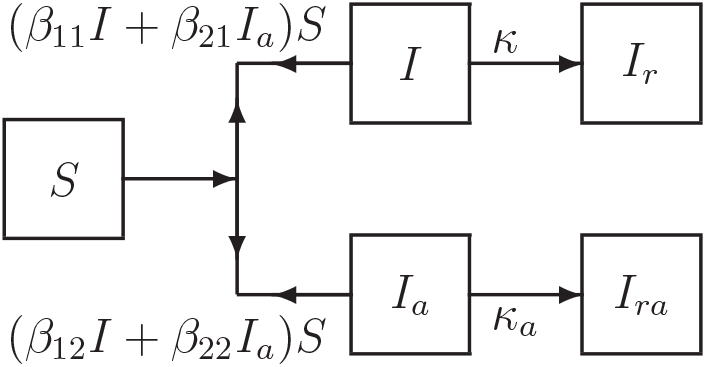
Flowchart of disease transmission among susceptible population *S*, current symptomatic infected individuals *I*, current asymptomatic infected individuals *I*_*a*_, recovered symptomatic infected individuals *I*_*r*_, recovered asymptomatic infected individuals *I*_*ra*_.

Assume that the dynamics of an epidemic can be described by *m* time intervals. At *l*th time interval, SARDDE (1) becomes the form:

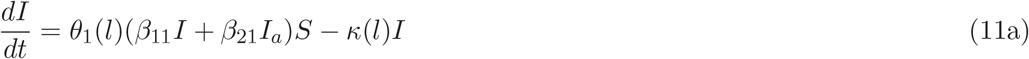

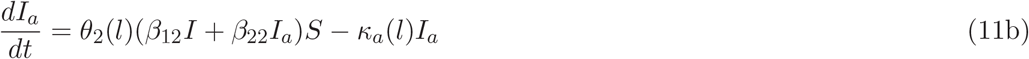

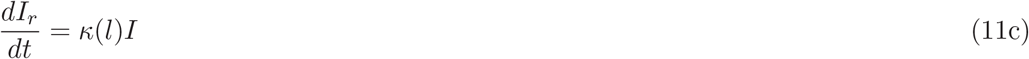

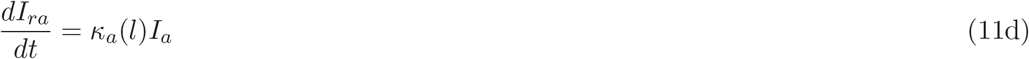

Then SARDDE (11) has a disease-free equilibrium:

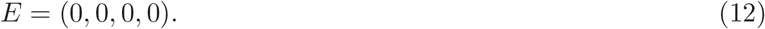

#### 2.3.2 Stability of disease-free equilibrium

The stability of SARDDE (11) is determined by the first two equations (11a) and (11b). Denote in (11a) and (11b):

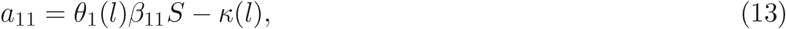

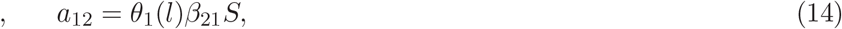

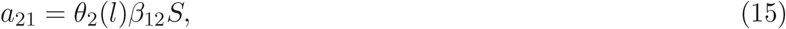

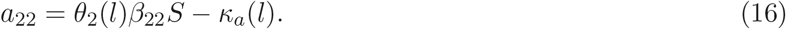

Then at the disease-free equilibrium of SARDDE(11), the Jacobian matrix of (11a) and (11b) has the form

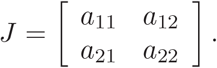

Solving the corresponding eigenequation obtains 2 eigenvalues:

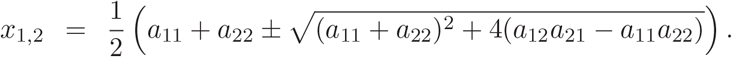

Therefore we obtain the following:

##### Theorem 3

*Suppose that a*_11_, *a*_12_, *a*_21_ *and a*_22_ *are defined by (13)-(16). Then the disease-free equilibrium E of SARDDE (11) is globally asymptotically stable if, and only if, the following inequalities hold:*

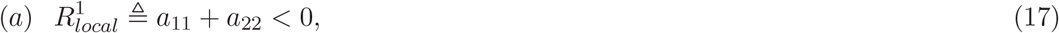

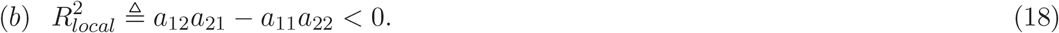

#### 2.3.3 The necessary condition of disease spreading

If an epidemic can occur, then

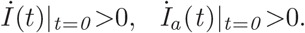

This implies that

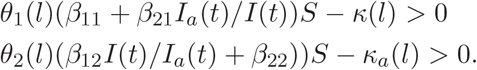

Solving the above inequalities gives the following

##### Theorem 4

*If SARDDE (11) satisfies the following inequalities*

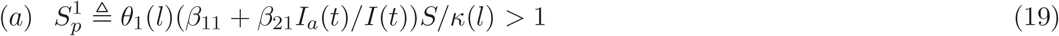

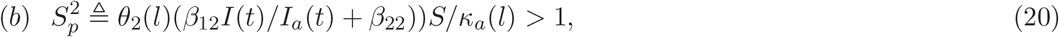

*Then a disease transmission will occur*.

## 3 Applications

### 3.1 Simulation and prediction of the first COVID-19 epidemic in Beijing

This Section will discuss the applications of Theorem 1, Theorem 2 and SARDDE (1) to simulate the real-word COVID-19 epidemic data from 19 January to 8 June 2020 in Beijing [17]. Numerical simulations and drawings are performed by using MATLAB software programs.

#### 3.1.1 Modeling and Simulation

Figures 3(a) and 3(b) show the first 55 days’ reported clinical data on the current confirmed infection cases, and the reported clinical data on recovered cases of the COVID-19 epidemic in Beijing [17]^1^. The number of the current symptomatic infected individuals is shown in Fig4(a) by circles. The numbers of the cumulative recovered symptomatic infected individuals, and the cumulative died infected individuals are shown in Fig4(b) by circles and stars respectively.

**Figure 3.**
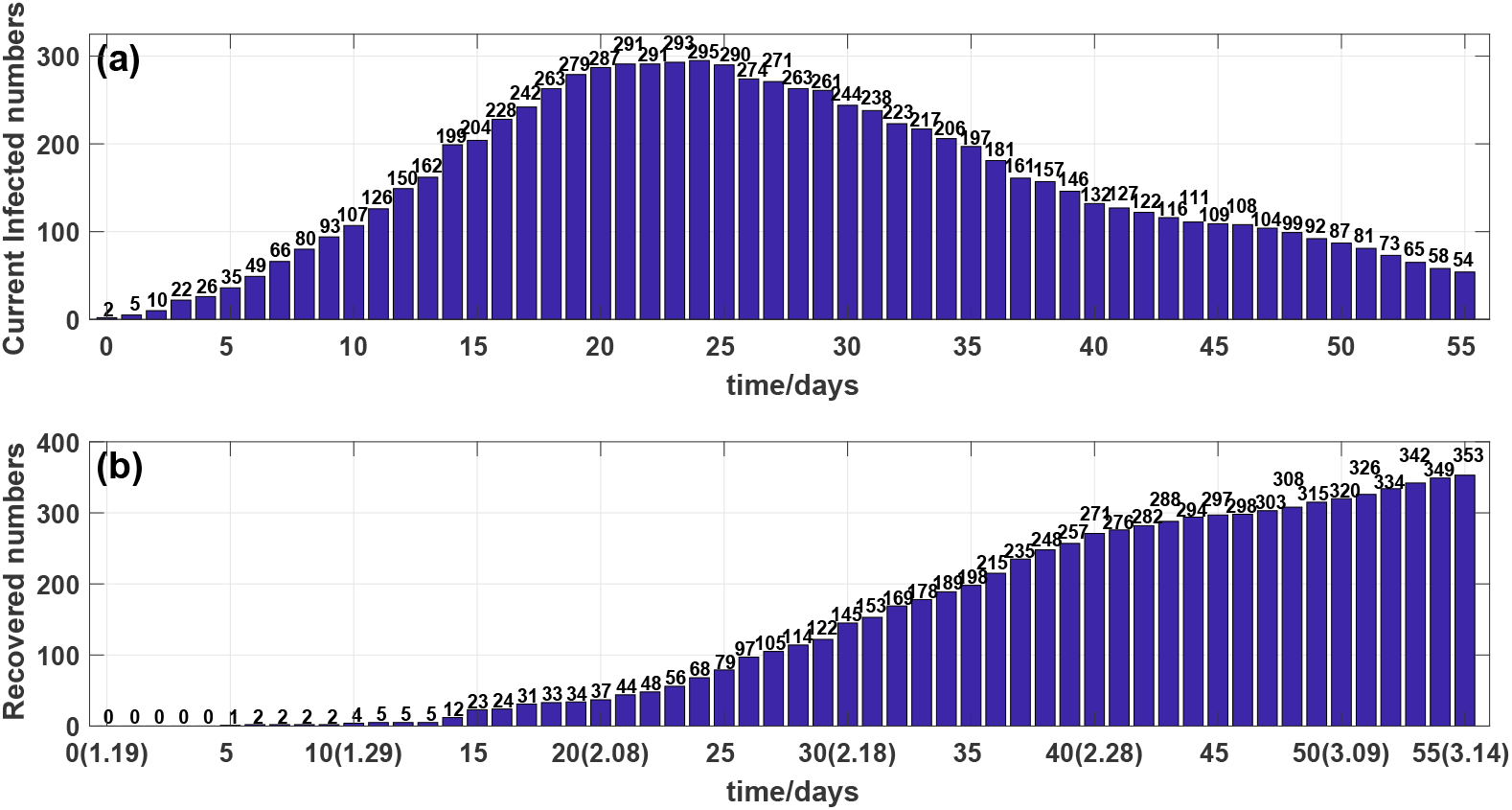
(a) Outcome of the number of the current infected individuals. (b) Outcome of the number of the cumulative recovered individuals.

The number of the current infected individuals was risen rapidly in the first 4 days (see Fig.3(a) and Fig. 4(a)). The number of the current infected individuals reached the highest 295 on day 24, February 12. Then after day 31, February 19, they declined rapidly (see Fig.3(a) and 4(a)).

**Figure 4.**
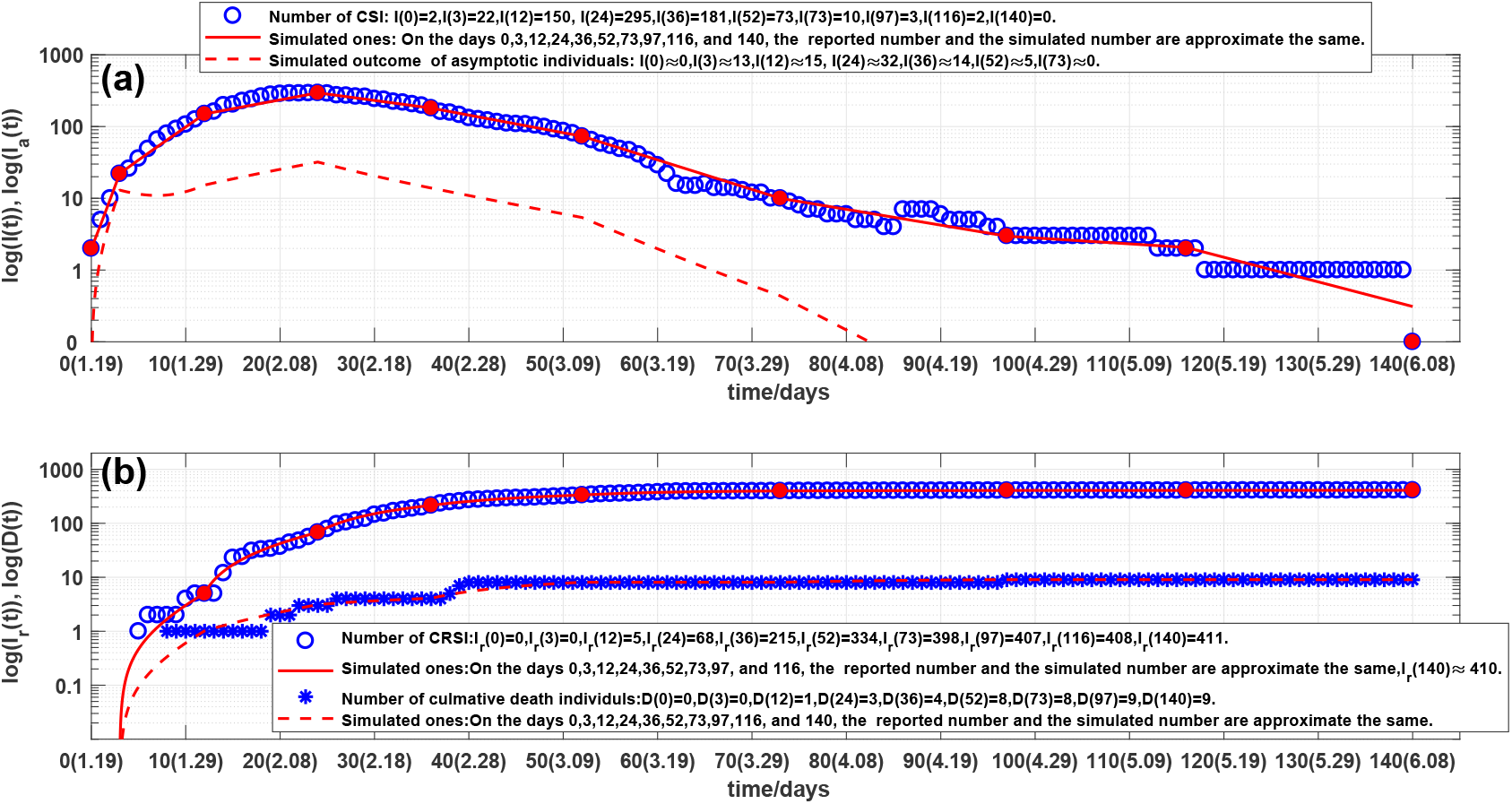
(a) Outcome of the number of the current symptomatic individuals (CSI), representing by circles. Solid lines and dash lines are outcomes of the corresponding simulations of SARDDE (1). (b) Outcomes of the numbers of the cumulative recovered symptomatic individuals (CRSI) and the cumulative died individuals (CDI), representing by circles and stars, respectively. Solid lines and dash lines are the corresponding simulations of SARDDE (1).

On day 86 (15 April), there were 3 Chaoyang district infected people coming back Beijing form foreign country which made calculated blocking rate rise. After day 97, the number of the current infected individuals were reducing continually (in piecewise linear way) until day 140 (8 June) reaching to zero.

Observe from Figs. 4(a) and 4(b) that the overall changes in the number of the current confirmed infections are not subject to the law of exponential changes, but the data can be approximated in good agreement with 9 straight lines in log scale (see Fig. 4). This phenomenon can be explained as different medical measures prevention and control strategies have been adopted over the different 9-time intervals. Therefore the *l* in SARDDE model (1) should be chosen as *l* = 1, 2, …, 9.

Denote

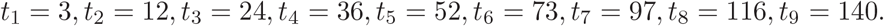

[*t*_*l*−1_, *t*_*l*_] is the *lth* time interval. Denote *I*_*c*_(*t*_*l*_) to be the number of the reported current symptomatic individuals, and Denote *I*_*cr*_(*t*_*l*_) to be the number of the reported current cumulative recovered symptomatic individuals. *D*_*c*_(*t*_*l*_) be the number of the reported current cumulative died individuals. Using the minimization error square criterion determines the *β*_*ij*_(*l*)^′^*s, κ*(*l*)^′^*s, κ*_*a*_(*l*)^′^*s*), *θ*_1_(*l*)^′^*s, θ*_2_(*l*)^′^*s*, and *α*(*l*)^′^*s*.

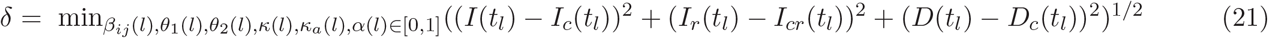

##### Remark

(1) One can assume that *S* = 1 because the effects of *S* can be deleted by calculated 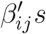. This makes the calculated 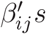 have general sense.

(2) In the previous version of this paper [18], *κ*(*l*), *α*(*l*) were determined via using the number of the recovered patients and died patients to divide the days of patients stayed in the hospital during the *lth* time interval, respectively. However It finds that with the helps of the formulas *κ*(*l*), and *α*(*l*) [18], we determine the search ranges of *κ*(*l*) and *α*(*l*) and can obtain better simulation results as those described later.

(3) The website [17] did not reported the data of asymptomatic infected individuals and announced only that there were no asymptomatic infected individuals after day 73 (April 1). Hence we cannot estimate the errors between the simulated *I*_*a*_(*l*) and *I*_*ra*_(*l*) with the reported ones.

(4) The search ranges of the infection rate 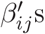 s are difficult to be determined. However if we know the number of current symptomatic infected individuals at day 0 and day T when no prevention and control measures had been implemented. Additional the number of current asymptomatic infected individuals was very small. Then we can estimate 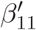 from equation (1a) as follows

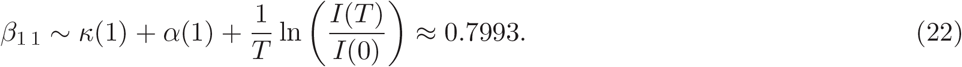

Using the real world data of the first COVID-19 epidemic in Beijing [17] selects following initial condition

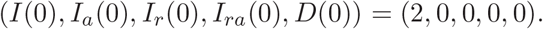

The calculated parameters are shown in Table 1. The corresponding simulation results of equation (1) are shown in Fig. 1 and Fig. 2. Observe that the simulation results of equation (1) were in good agreement with the data of the COVID-19 epidemics. At the end points (see solid dots in Figs. 1 and 2) of the 9 investigated time-interval [*t*_*l*−1_, *t*_*l*_]^′^*s*, the simulated numbers and the actual reported numbers were approximate the same∼ errors were less than one, except on day 140, the numbers of the simulated and the reported cumulative recovered infected individuals was one difference. (See Figs. 4(a) and 4(b) and legends in them).

**Table 1.**
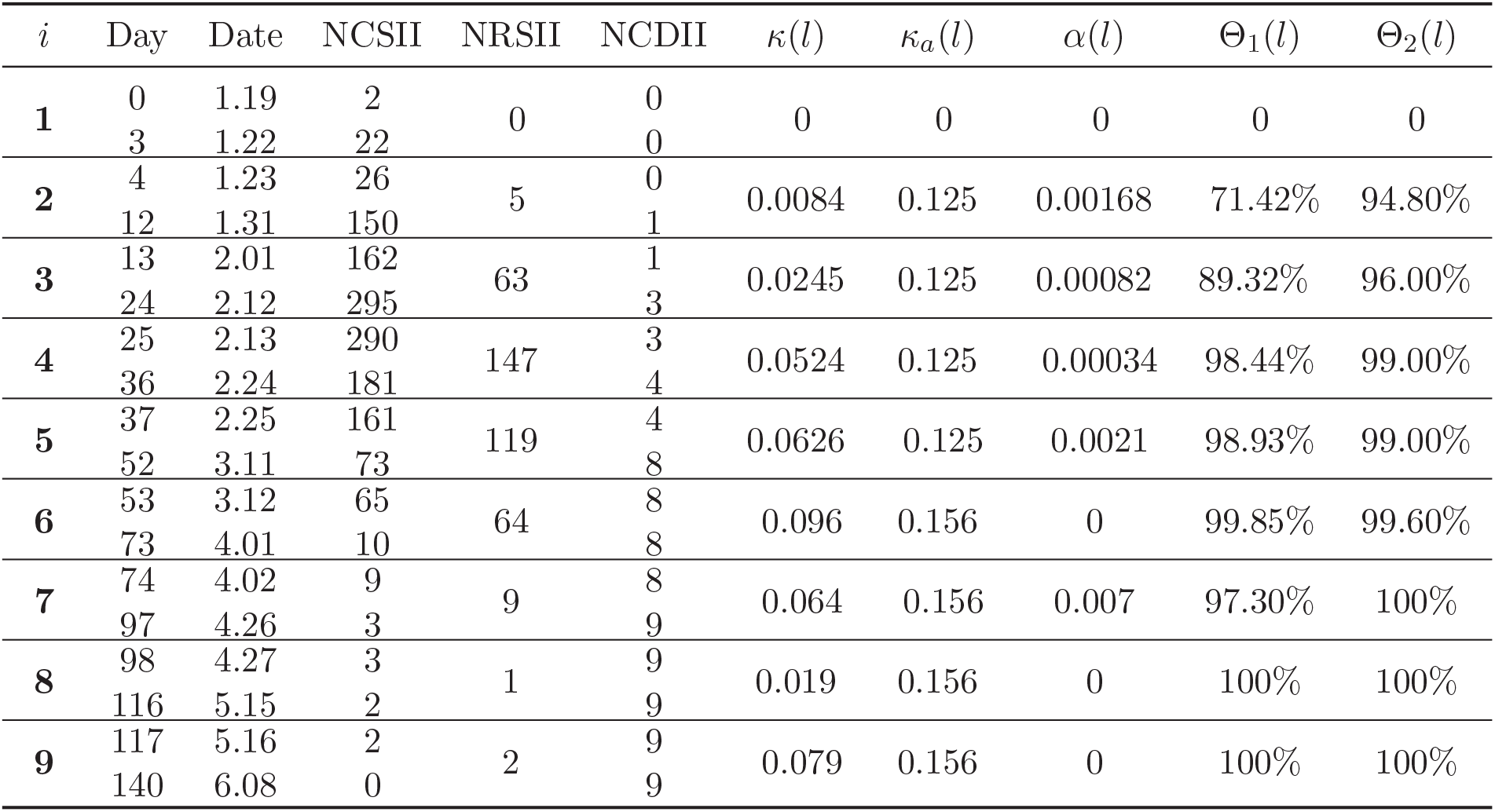
The data of the first COVID-19 epidemic in Beijing on different days and corresponding calculated SARDDE parameters. Where NCSII and NCDI represent the numbers [17] of the current symptomatic infected individuals and the cumulative died individuals, respectively; NRSII the number [17] of the recovered symptomatic infected individuals over the *lth* time interval.*β*_11_ = 0.75454, *β*_21_ = 0.1119, *β*_12_ = 0.48757, *β*_22_= 0.07993.

| $i$ | Day | Date | NCSII | NRSII | NCDII | $\kappa(l)$ | $\kappa_a(l)$ | $\alpha(l)$ | $\Theta_1(l)$ | $\Theta_2(l)$ |
| --- | --- | --- | --- | --- | --- | --- | --- | --- | --- | --- |
| **1** | 0 | 1.19 | 2 | 0 | 0 | 0 | 0 | 0 | 0 | 0 |
|  | 3 | 1.22 | 22 |  | 0 |  |  |  |  |  |
| **2** | 4 | 1.23 | 26 | 5 | 0 | 0.0084 | 0.125 | 0.00168 | 71.42% | 94.80% |
|  | 12 | 1.31 | 150 |  | 1 |  |  |  |  |  |
| **3** | 13 | 2.01 | 162 | 63 | 1 | 0.0245 | 0.125 | 0.00082 | 89.32% | 96.00% |
|  | 24 | 2.12 | 295 |  | 3 |  |  |  |  |  |
| **4** | 25 | 2.13 | 290 | 147 | 3 | 0.0524 | 0.125 | 0.00034 | 98.44% | 99.00% |
|  | 36 | 2.24 | 181 |  | 4 |  |  |  |  |  |
| **5** | 37 | 2.25 | 161 | 119 | 4 | 0.0626 | 0.125 | 0.0021 | 98.93% | 99.00% |
|  | 52 | 3.11 | 73 |  | 8 |  |  |  |  |  |
| **6** | 53 | 3.12 | 65 | 64 | 8 | 0.096 | 0.156 | 0 | 99.85% | 99.60% |
|  | 73 | 4.01 | 10 |  | 8 |  |  |  |  |  |
| **7** | 74 | 4.02 | 9 | 9 | 8 | 0.064 | 0.156 | 0.007 | 97.30% | 100% |
|  | 97 | 4.26 | 3 |  | 9 |  |  |  |  |  |
| **8** | 98 | 4.27 | 3 | 1 | 9 | 0.019 | 0.156 | 0 | 100% | 100% |
|  | 116 | 5.15 | 2 |  | 9 |  |  |  |  |  |
| **9** | 117 | 5.16 | 2 | 2 | 9 | 0.079 | 0.156 | 0 | 100% | 100% |
|  | 140 | 6.08 | 0 |  | 9 |  |  |  |  |  |

#### 3.1.2 Discussions

On the end points of the 9 investigated time-interval [*t*_*l*−1_, *t*_*l*_]^′^*s*, that is on day 0, day 3, day 12, day 24, day 30, day 243, day 73, day 97, day 116, and day 140, we obtain the following results (also see Figs. 4).

1. On the end point days, the reported and simulated current symptomatic individuals are are approximate the same.
2. On day 140, the numbers of the reported and simulated cumulative recovered individuals were one difference. On the other end point days, the numbers were the same.
3. On the end point days, the numbers of the reported and simulated cumulative died individuals are approximate the same.
4. There is no information on the current symptomatic infected and recovered symptomatic infected individuals. But it has reported that after the 73 day, April 1, there is no symptomatic infected individuals until day 143, June 11 [17]. Our simulation results shows that on day 73, the number of the simulated current symptomatic infected individuals was less than one (≈ 0.43612), the cumulative asymptomatic infected individuals discharged from observations were about 104 individuals, which seem to explain the actual missing report asymptomatic infected individual data.
5. Computed results (see Table 1) of the transmission rates 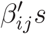 show that the ratio of the transmission rates of asymptomatic and symptomatic individuals infecting susceptible population to become symptomatic individuals is about 0.159 (*β*_21_:*β*_11_). It suggests that the asymptomatic individuals cause lesser symptomatic spread than the symptomatic individuals do.
6. The computed results (see Table 1) also show that the ratios of the transmission rates of the asymptomatic and the symptomatic individuals infecting susceptible population to become the asymptomatic and the symptomatic individuals are about 0.646 (*β*_12_ : *β*_11_) and 0.667 (*β*_22_ : *β*_21_), respectively. It suggests that both the symptomatic and the asymptomatic individuals cause lesser asymptomatic spreads than symptomatic spreads.
7. The criteria (7) and (8) of the asymptotical stability of the disease-free equilibrium of SARDDE model (1) over the 8-time intervals are shown in the 5th ∼ 8th columns of Table 2. It is shown that until the blocking rates (Θ_1_, Θ_2_) reach about (98.44%, 99%), the disease-free equilibrium becomes globally asymptotical stability. The conditions (9) and (10) of disease spreading are listed in the last two columns in Table 2. It shows also that if the blocking rates (*θ*_1_, *θ*_2_) reach about (98.44%, 99%), the spreading of COVID-19 epidemic can be blocked. Table 2 suggests also that 90% and 96% blocking rates to symptomatic infections and asymptomatic infections cannot prevent spreads to transmissions of symptomatic infections and asymptomatic infections.

**Table 2.**
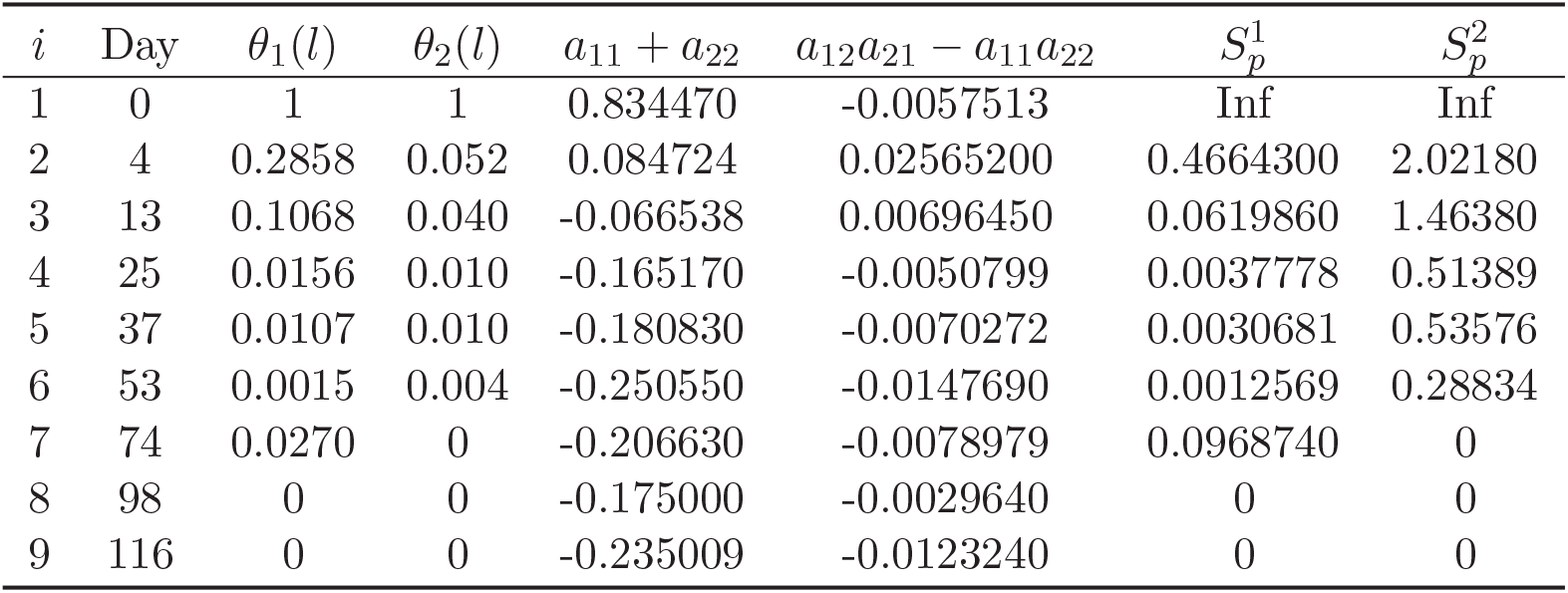
Criteria of the asymptotical stability and disease spreading of disease-free equilibrium of SARDDE (1) over 9-time intervals.

| $i$ | Day | $\theta_1(l)$ | $\theta_2(l)$ | $a_{11} + a_{22}$ | $a_{12}a_{21} - a_{11}a_{22}$ | $S_p^1$ | $S_p^2$ |
| --- | --- | --- | --- | --- | --- | --- | --- |
| 1 | 0 | 1 | 1 | 0.834470 | -0.0057513 | Inf | Inf |
| 2 | 4 | 0.2858 | 0.052 | 0.084724 | 0.02565200 | 0.4664300 | 2.02180 |
| 3 | 13 | 0.1068 | 0.040 | -0.066538 | 0.00696450 | 0.0619860 | 1.46380 |
| 4 | 25 | 0.0156 | 0.010 | -0.165170 | -0.0050799 | 0.0037778 | 0.51389 |
| 5 | 37 | 0.0107 | 0.010 | -0.180830 | -0.0070272 | 0.0030681 | 0.53576 |
| 6 | 53 | 0.0015 | 0.004 | -0.250550 | -0.0147690 | 0.0012569 | 0.28834 |
| 7 | 74 | 0.0270 | 0 | -0.206630 | -0.0078979 | 0.0968740 | 0 |
| 8 | 98 | 0 | 0 | -0.175000 | -0.0029640 | 0 | 0 |
| 9 | 116 | 0 | 0 | -0.235009 | -0.0123240 | 0 | 0 |

#### 3.1.3 Virtual Simulations

Now assume that after day 24, 12 February, it still keeps the blocking rates (Θ_1_(3), Θ_2_(3)) ≈ (89.93%, 96%), the recoverr rates (*κ*(3), *κ*_*a*_(3)), and the died rate *α*(3) until day 140, 8 June. The simulation results of SARDDE (1) are shown in Figs 5(a) and 5(b). Observe that the numbers of current symptomatic and the asymptomatic infected individuals reach about 208958 and 22847, respectively. The numbers of cumulative recovered symptomatic and died individuals reach about 90436 and 3028, respectively.

**Figure 5.**
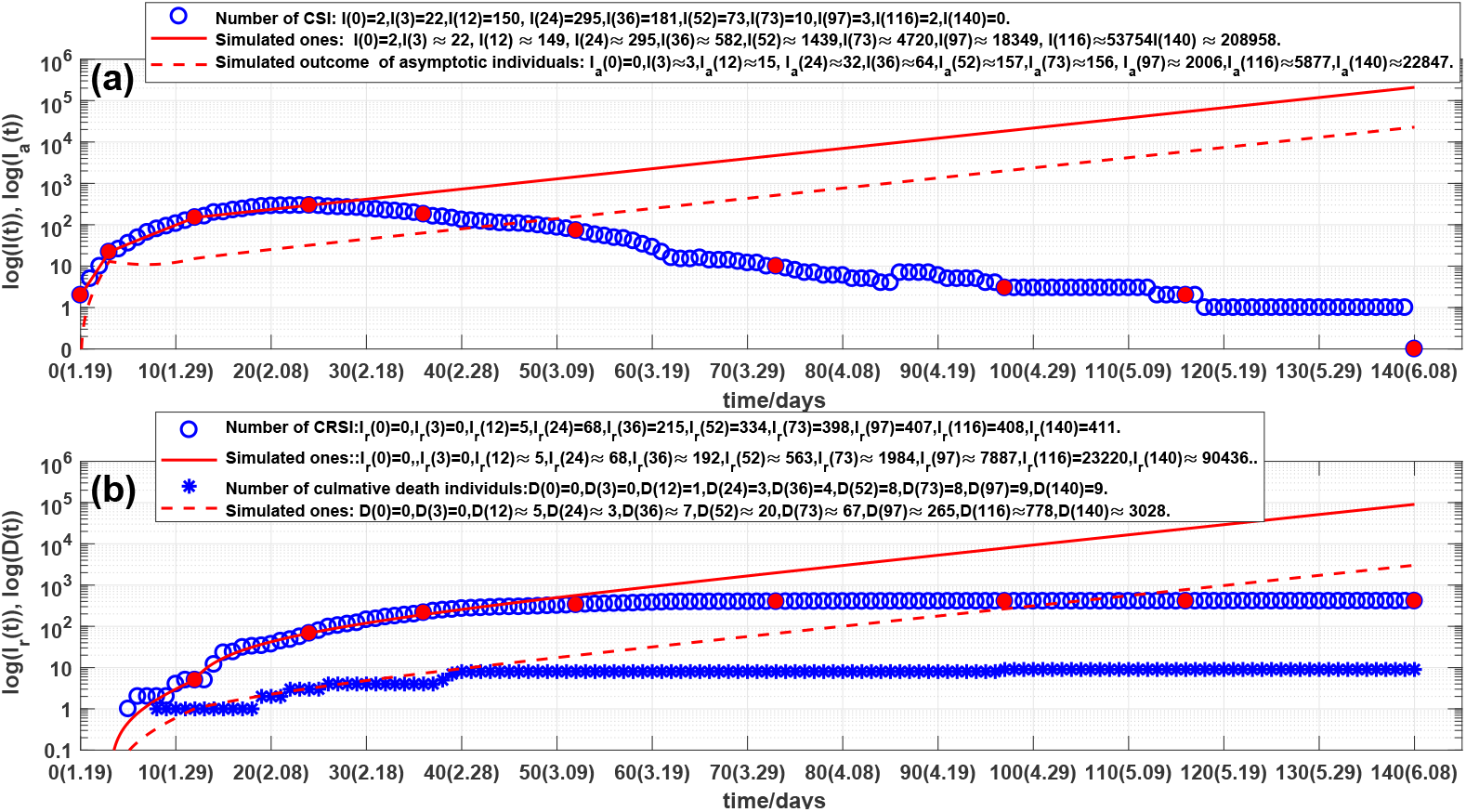
Virtual simulations: (a) Outcome of the number of the current symptomatic individuals (CSI), representing by circles. Solid lines and dash lines are outcomes of the corresponding simulations of SARDDE (1). (b) Outcomes of the numbers of the cumulative recovered symptomatic individuals (CRSI) and the cumulative died individuals (CDI), representing by circles and stars, respectively. Solid lines and dash lines are the corresponding simulations of SARDDE (1).

Furthermore assume that after day 52, 11 March, it still keeps the blocking rates (Θ_1_(5), Θ_2_(4)) ≈ (98.93%, 99%), the recovery rates (*κ*(5), *κ*_*a*_(5)), and the died rate *α*(5) until day 140, 8 June. The simulation results of SARDDE (1) are shown in Figs 6(a) and 6(b). Observe that the numbers of current symptomatic and asymptomatic infected individuals are about one and zero, respectively; the numbers of cumulative recovered symptomatic and died individuals are about 415 and 11, respectively. The results suggest that using the data before day 52 (about 17 days after the turning point) can estimate well the following outcome of the first COVID-19 epidemic in Beijing.

**Figure 6.**
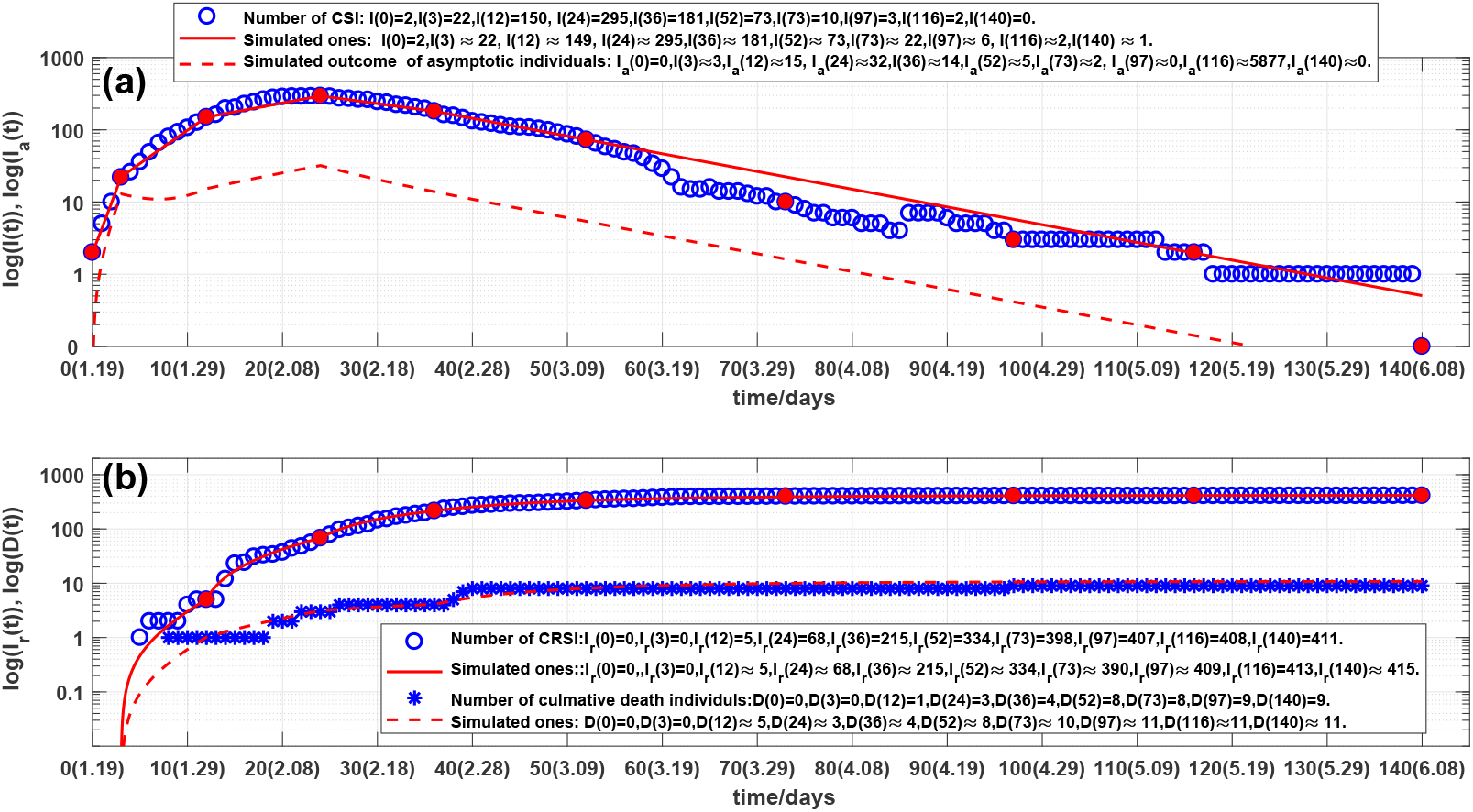
Virtual simulations: (a) Outcome of the number of the current symptomatic individuals (CSI), representing by circles. Solid lines and dash lines are outcomes of the corresponding simulations of SARDDE (1). (b) Outcomes of the numbers of the cumulative recovered symptomatic individuals (CRSI) and the cumulative died individuals (CDI), representing by circles and stars, respectively. Solid lines and dash lines are the corresponding simulations of SARDDE (1).

In summary, SARDDE (1) can simulate the outcomes of the first COVID-19 epidemic in Beijing. The calculated equation parameters can help us to understand and explain the mechanism of epidemic diseases and control strategies for the event of the practical epidemic.

### 3.2 Simulation and prediction of the second COVID-19 epidemic in Beijing

This Section will discuss the applications of Theorem 3, Theorem 4 and SARDDE (11) to simulate the real-word COVID-19 epidemic data from 11 June to 6 August 2020 in Beijing [17]. This event of the second epidemic in Beijing provides a valuable example of accurate preventing and controlling strategies and excellent clinical treatments. The data of the symptomatic and asymptomatic infected and recovered individuals provide a footstone to modeling the event.

#### 3.2.1 Modeling and Simulation

Figure 7(a) show that the reported data on the current confirmed symptomatic infection cases. Figure 7(b) show that the reported data on the cumulative recovered symptomatic infection cases. Figure 8(a) show that the reported data on the current confirmed asymptomatic infection cases. Figure 8(b) show that the reported data on the cumulative recovered asymptomatic infection cases. All data set were edited from [17].

**Figure 7.**
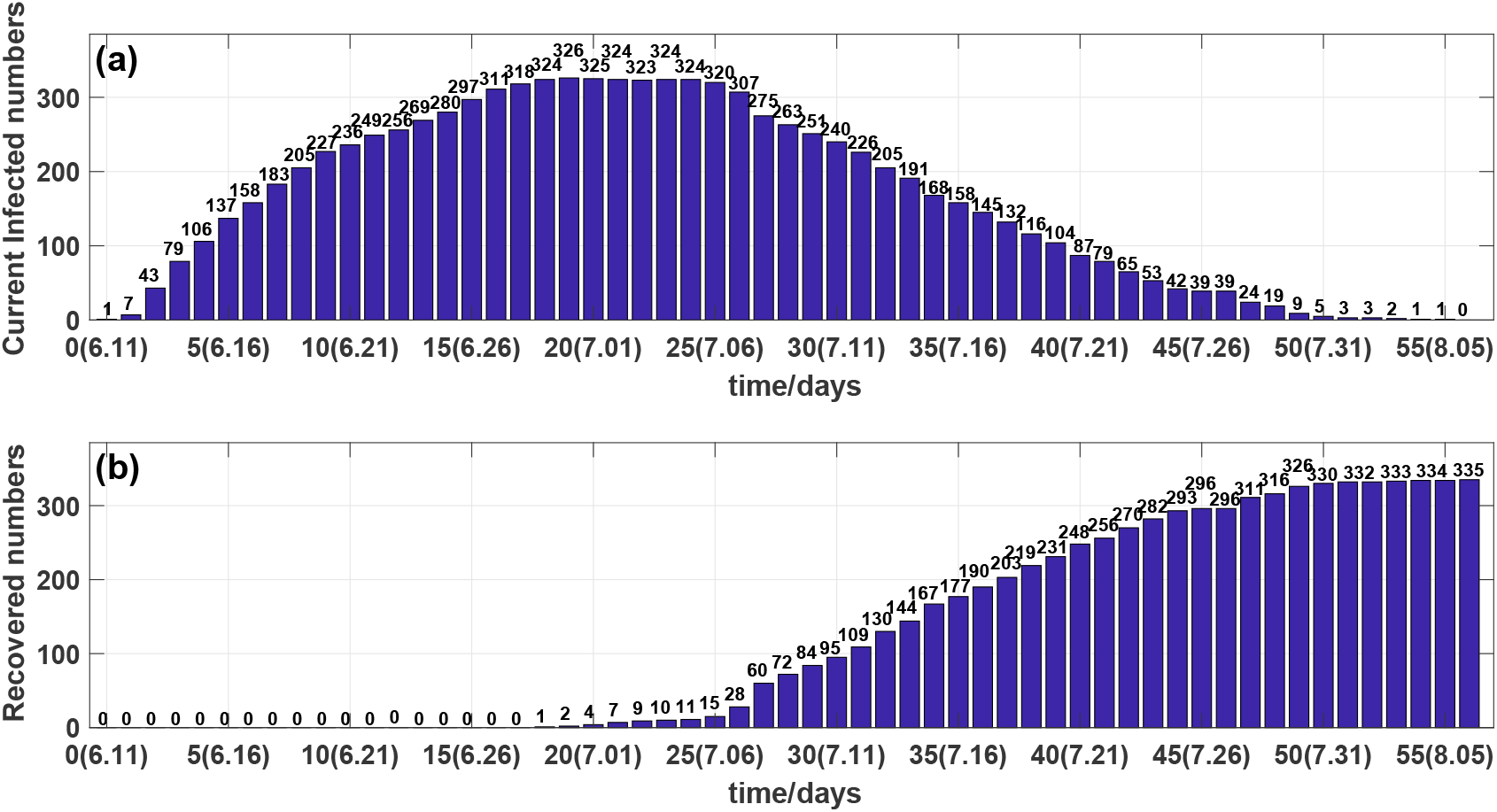
(a) Outcome of the number of the current symptomatic infected individuals. (b) Outcome of the number of the cumulative recovered symptomatic infected individuals.

**Figure 8.**
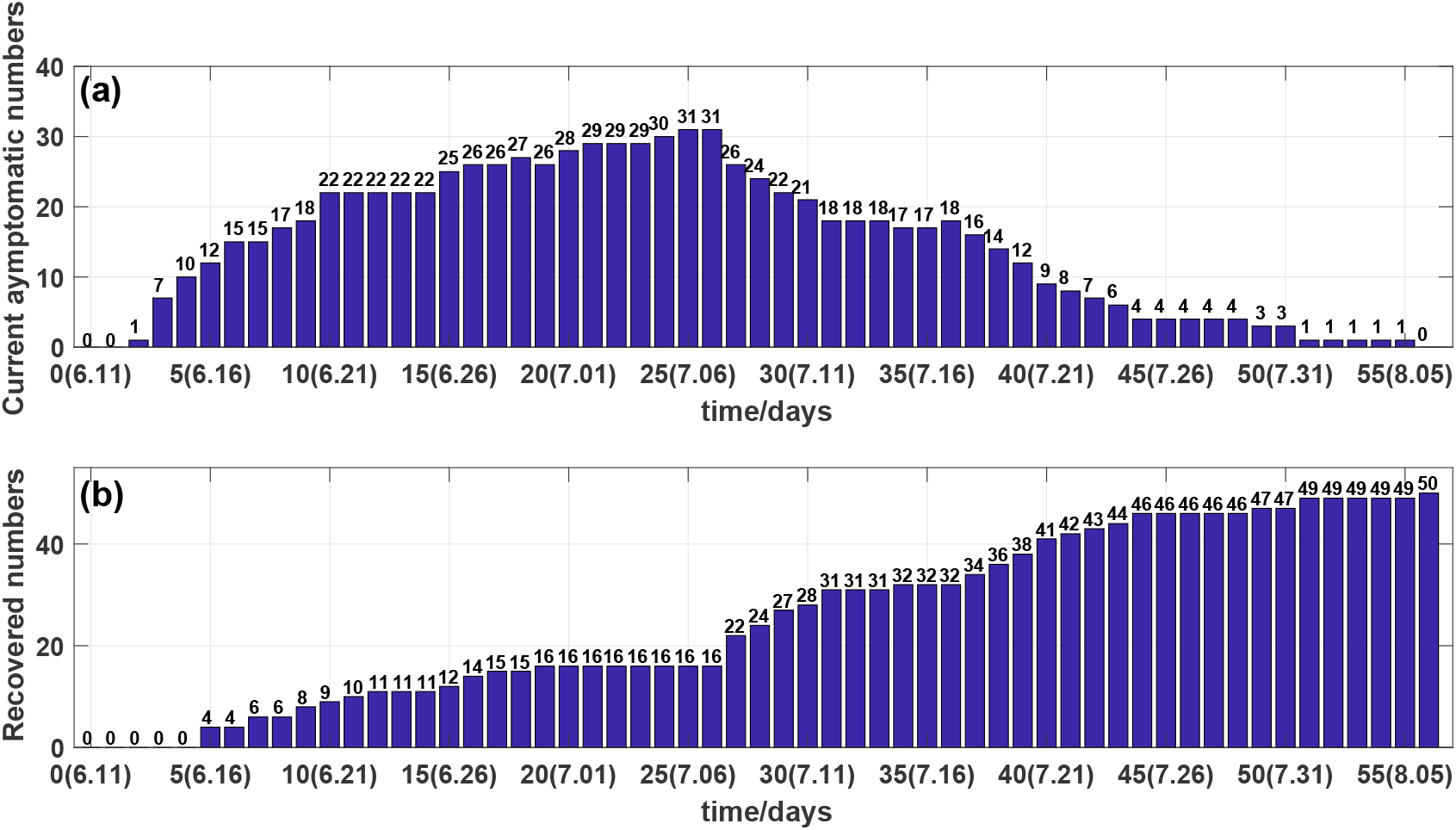
(a) Outcome of the number of the current asymptomatic infected individuals. (b) Outcome of the number of the cumulative recovered asymptotic infected individuals.

The evolution of the current symptomatic infected individuals, and the current asymptomatic infected individuals are shown in Fig. 9(a) by circles and diamonds, respectively. The evolution of the cumulative recovered symptomatic infected individuals, and the cumulative recovered asymptomatic infected individuals are shown in Fig. 9(b) by circles and diamonds, respectively.

**Figure 9.**
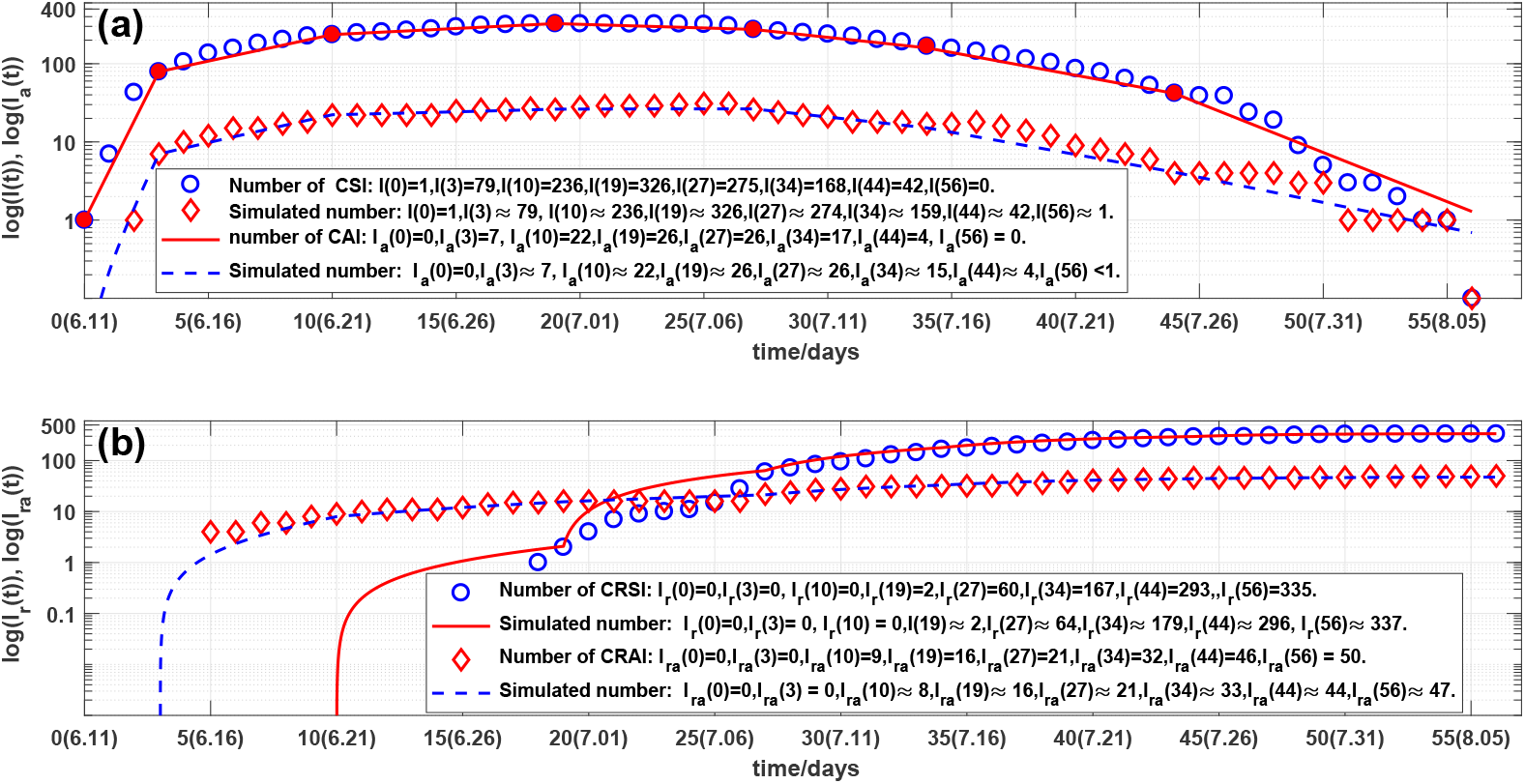
Outcomes of the numbers of: (a) the current symptomatic individuals (CSI) and the current asymptomatic individuals (CAI), representing by circles and diamonds. Solid lines and dash lines are the corresponding simulations of SARDDE (11). Outcomes of the numbers of: (b) the cumulative recovered symptomatic individuals (CRSI) and the cumulative recovered asymptomatic individuals (CRAI), representing by circles and diamonds, respectively. Solid lines and dash lines are the corresponding simulations of SARDDE (11).

Observe from Fig.9 that the overall changes in the number of the current confirmed infections are not subject to the law of exponential changes, but the data can be approximated in good agreement with 8 straight lines in log scale (see Fig.9). This phenomenon can be explained as different medical measures and prevention and control strategies have been adopted over different 8-time intervals. Therefore the *l* in SARDDE (11) satisfies *i* = 1, 2, …, 8.

Denote

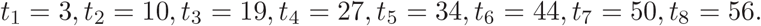

Denote *I*_*c*_(*t*_*l*_) to be the number of the reported current symptomatic individuals, and *I*_*a*_(*t*_*l*_) be the number of the reported current asymptomatic individuals charged in medical medical observations. Denote *I*_*cr*_(*t*_*l*_) to be the number of the reported current cumulative recovered symptomatic individuals. and *I*_*cra*_(*t*_*l*_) be the number of the reported current cumulative asymptomatic individuals discharged from medical medical observations.

Using the minimization error square criterion (23) determines the *β*_*ij*_(*l*)^′^*s, κ*(*l*)^′^*s, κ*_*a*_(*l*)^′^*s*), *θ*_1_(*l*)^′^*s* and *θ*_2_(*l*)^′^*s*.

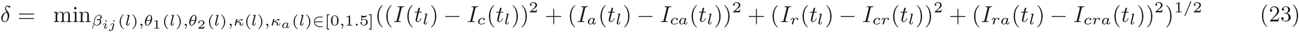

The calculated results are shown in Table 3. The corresponding simulation results of SARDDE (11) are shown in Fig. 9(a) and 9(b). Observe that the simulation results of model (11) describe well the dynamics of the second COVID-19 epidemic in Beijing.

**Table 3.**
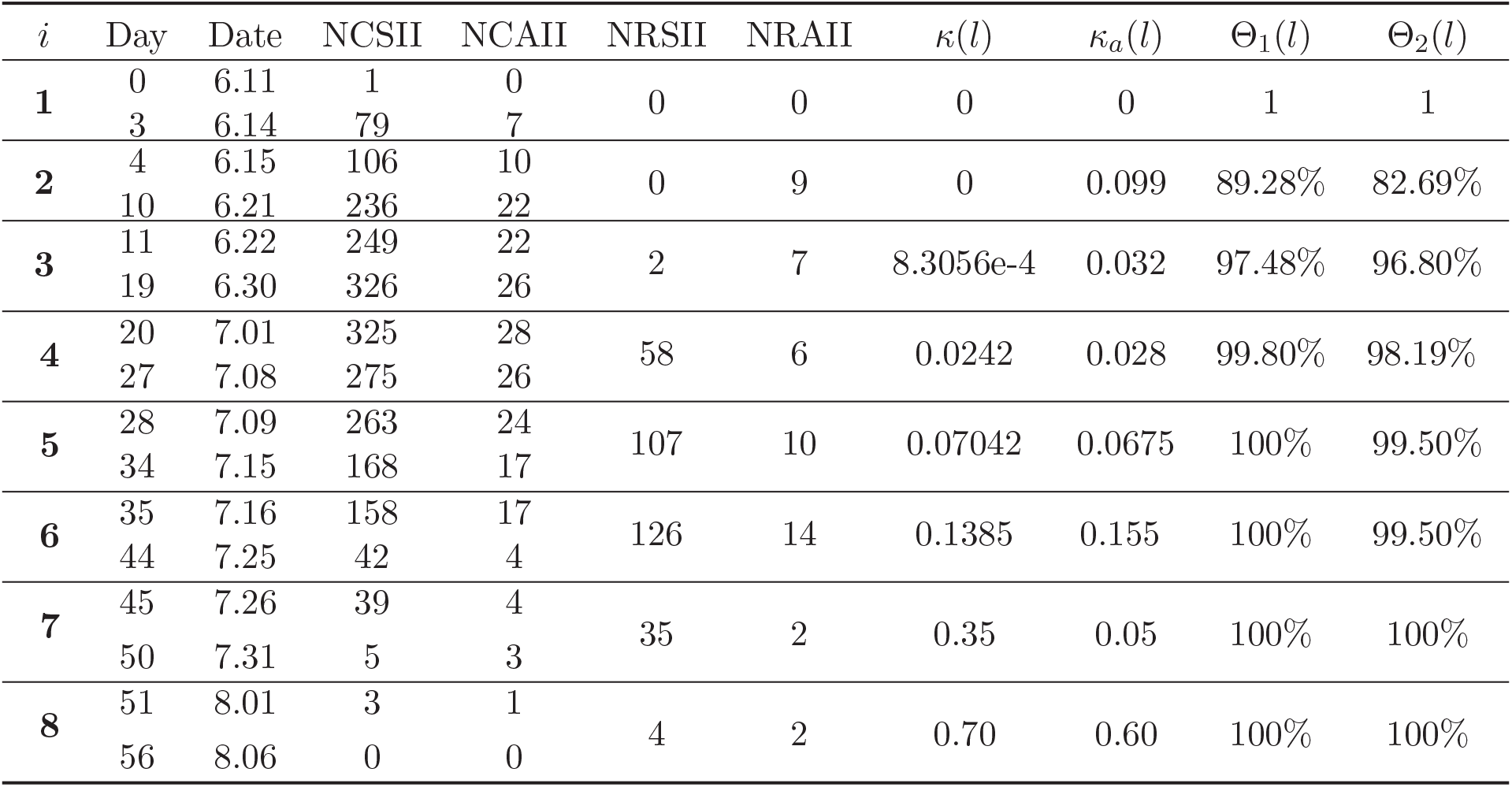
The data of the second wave COVID-19 epidemics on different days and the corresponding calculated parameters of SARDDE model (11). Where NCSII and NCAII represent the numbers of the current symptomatic infected individuals and the current asymptomatic infected individuals, respectively; NRSII and NRAII represent the numbers of the cumulative recovered symptomatic infected individuals and the asymptomatic infected individuals over the *i*th interval. *β*_11_ = 1.4521, *β*_21_ = 0.072824, *β*_12_ = 0.072824, *β*_22_ = 0.72824.

##### Remark

(1) Similar the description in Section 3.1.1, we have

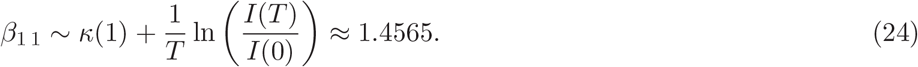

Using the practical data of the second COVUD-19 epidemic (also see the second line in Table 3) selects following initial condition

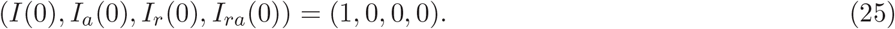

The simulations of SARDDE (11) with the above model parameters are shown in Figs. 9(a) and 9(b). Observe that the simulation results are in good agreement with the reported clinical data On the nine time interval endpoint days, the errors between the reported data and the simulated data were less than one (see the solid and dash lines and legends in Figs. 9(a) and 9(b)).

#### 3.2.2 Discussions

On the end points of the 8 investigated time-interval [*t*_*l*−1_, *t*_*l*_]^′^*s*, that is on day 0, day 3, day 10, day 19, day 27, day 34, day 50, and day 55, we obtain the following results (also see Figs. 9 and legends).

1. On the end point days, the numbers of the reported and simulated current symptomatic individuals were approximate the same.
2. On the end point days, the numbers of the reported and simulated current asymptomatic individuals were approximate the same.
3. On the end point days, the numbers of the reported and simulated cumulative recovered symptomatic individuals were approximate the same.
4. On the end point days, the numbers of the reported and simulated cumulative recovered asymptomatic individuals were approximate the same.
5. On the end point days, the numbers of the reported and simulated cumulative died individuals are approximate the same.
6. Computed results (see Table 3) show that the ratio of the transmission rates of the asymptomatic and the symptomatic individuals infecting susceptible population to become the symptomatic individuals is about 5% (*β*_21_ : *β*_11_). It suggests that the asymptomatic individuals cause lesser symptomatic spread than the symptomatic individuals do.
7. Computed results (see Table 3) also show that the ratios of the transmission rates of the asymptomatic and symptomatic individuals infecting susceptible population to become the asymptomatic and symptomatic individuals are about 5% (*β*_12_:*β*_11_). It suggests that the symptomatic individuals cause lesser asymptomatic spread than symptomatic spread.
8. The criteria of the stability of the disease-free equilibrium of SARDDE (11) on 8-time intervals are shown in Table 4. It shows that the blocking rates reach about (97.47%, 96.6%) cannot prevent the spread of the second COVID-19 epidemic in Beijing; the blocking rates (Θ_1_, Θ_2_) reach about (99.80%, 98.19%), the disease-free equilibrium becomes globally asymptotical stable.

**Table 4.** Criteria of the asymptotical stability and disease spreading of disease-free equilibrium of SARDDE (11) over 8-time intervals.

| $i$ | Day | $\theta_1(l)$ | $\theta_2(l)$ | $a_{11} + a_{22}$ | $a_{12}a_{21} - a_{11}a_{22}$ | $S_p^1$ | $S_p^2$ |
| --- | --- | --- | --- | --- | --- | --- | --- |
| 1 | 3 | 1 | 1 | 1.580300 | -0.00402600 | $+\infty$ | $+\infty$ |
| 2 | 10 | 0.1072 | 0.1731 | 0.169410 | -0.00402650 | $+\infty$ | 2.629300 |
| 3 | 19 | 0.0252 | 0.0320 | 0.038992 | 0.00031806 | 0.4534600 | 1.637200 |
| 4 | 27 | 0.0020 | 0.0181 | -0.021039 | -0.00031532 | 0.0014667 | 0.965280 |
| 5 | 34 | 0 | 0.0050 | -0.070420 | -0.00449690 | 0 | 0.106690 |
| 6 | 44 | 0 | 0.0050 | -0.138500 | -0.02096300 | 0 | 0.048657 |
| 7 | 50 | 0 | 0 | -0.350000 | -0.01750000 | 0 | 0 |
| 8 | 56 | 0 | 0 | -0.700000 | -0.42000000 | 0 | 0 |

#### 3.2.3 Virtual Simulation

Now assume that it keeps still the blocking rates (Θ_1_(3), Θ_2_(3)) ≈ (97.47%, 96.6%) and the cure rates (*κ*(3), *κ*_*a*_(3)) until day 56, 6 August. The simulation results of SARDDE (11) are shown in Fig.10. Observe that on day 56, the numbers of the current symptomatic and asymptomatic infected individuals reach about 1230 and 71, respectively; The numbers of the cumulative recovered symptomatic and asymptomatic infected individuals reach 23 and 67, respectively. If kept the above equation parameters until day 140, the numbers of the current symptomatic and asymptomatic infected individuals reach about 25017 and 1312, respectively; the numbers of the cumulative recovered symptomatic and asymptomatic infected individuals reach 574 and 1190, respectively.

**Figure 10.**
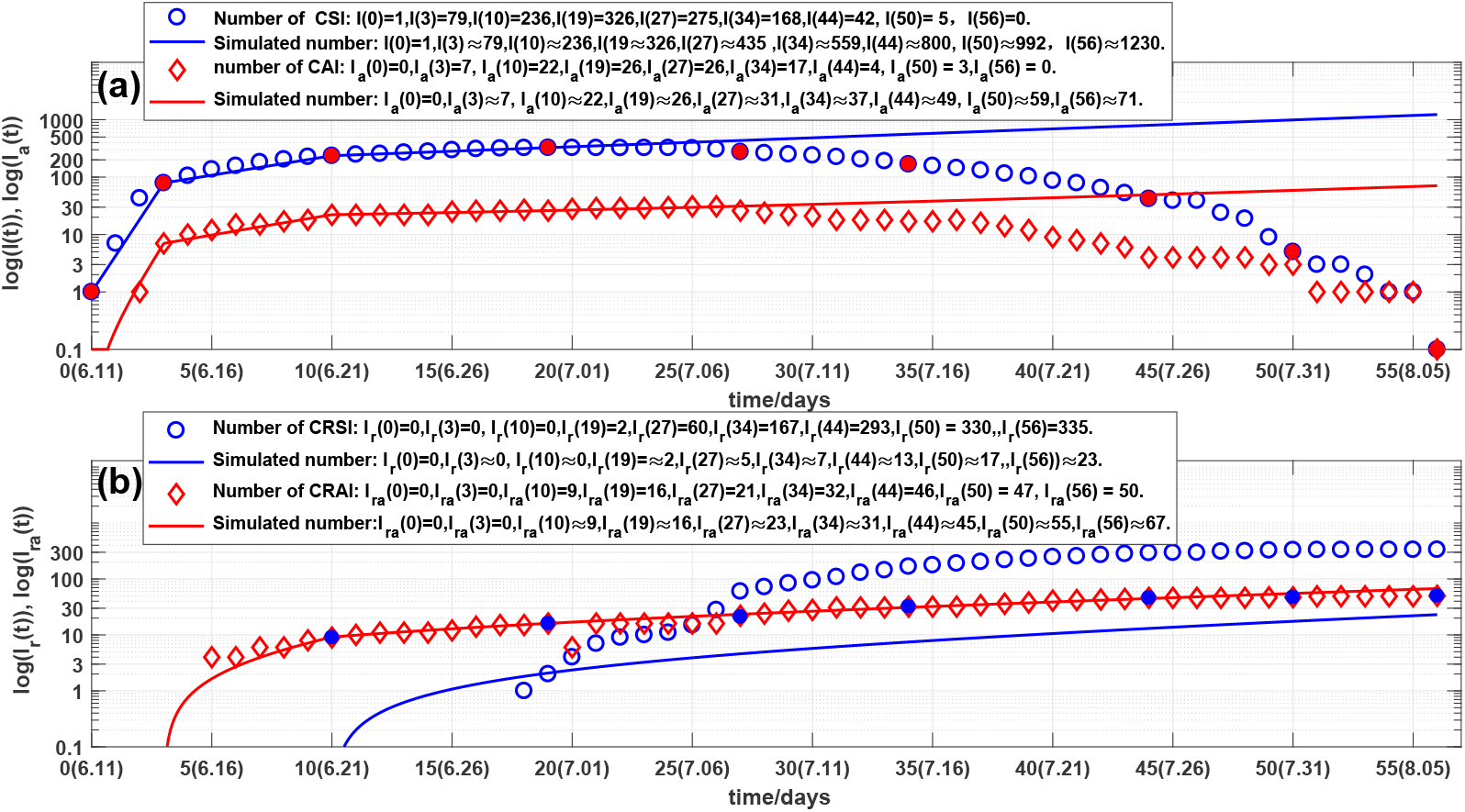
Virtual simulations. Outcomes of the numbers of: (a) the current symptomatic individuals (CSI) and the current asymptomatic individuals (CAI), representing by circles and diamonds. Solid lines and dash lines are the corresponding simulations of SARDDE (11). Outcomes of the numbers of: (b) the cumulative recovered symptomatic individuals (CRSI) and the cumulative recovered asymptomatic individuals (CRAI), representing by circles and diamonds, respectively. Solid and dash lines are the corresponding simulations of SARDDE (11).

Furthermore assume that after the day 34th, 14 July, it still keeps the blocking rates (Θ_1_(5), Θ_2_(5)), the cure rates (*κ*(5), *κ*_*a*_(5)) until day 56, 6 August. The simulation results of SARDDE (11) are shown in Figs. 11(a) and 11(b). Observe that on day 56, the numbers of the current symptomatic and asymptomatic infected individuals reach about 36 and 5, respectively. The numbers of the cumulative recovered symptomatic and the asymptomatic individuals are about 300 and 46, respectively. The results suggest that using the data before the day 34 (about two weeks after the turning point) can approximately estimate the following outcome of the second COVID-19 epidemic in Beijing.

**Figure 11.**
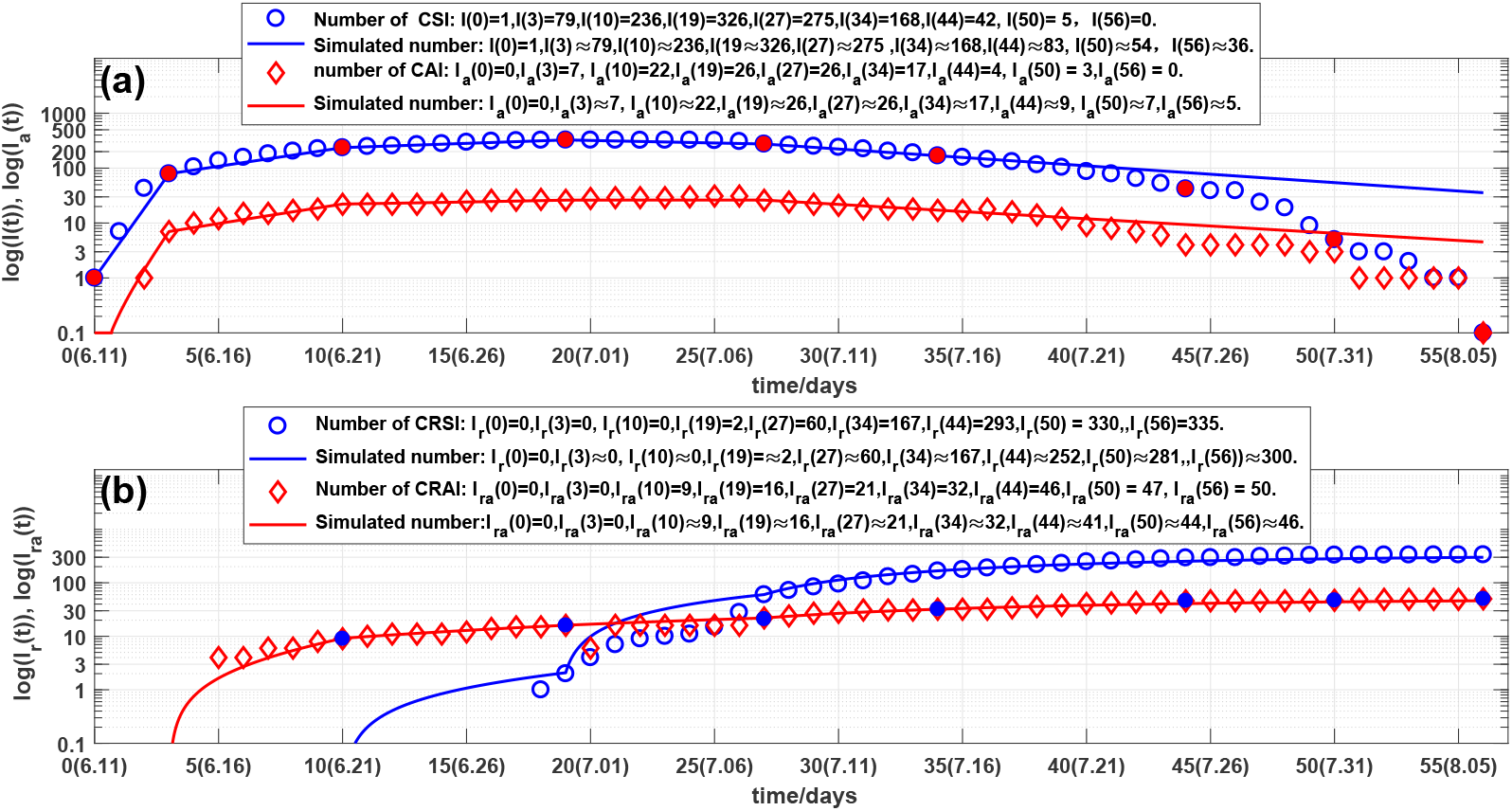
Virtual simulations. Outcomes of the numbers of: (a) the current symptomatic individuals (CSI) and the current asymptomatic individuals (CAI), representing by circles and diamonds. Solid lines and dash lines are the corresponding simulations of SARDDE (11). Outcomes of the numbers of: (b) the cumulative recovered symptomatic individuals (CRSI) and the cumulative recovered asymptomatic individuals (CRAI), representing by circles and diamonds, respectively. Solid and dash lines are the corresponding simulations of SARDDE (11).

### 3.3 Comparing

The model main parameters of the first and second COVID-19 in Beijing and related data are listed in Table 5. It follows:

**Table 5.**
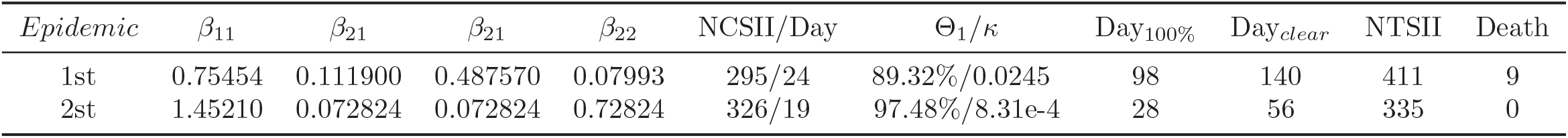
Model parameters and related data of the first and second COVID-19 epidemics in Beijing.

- The transmission rate *β*_11_ of the symptomatic infections caused by the symptomatic individuals in the second Beijing epidemic is much higher than the one in the first Beijing epidemic (1.4521:0.75454).
- The numbers of the highest hospitalized symptomatic individuals (NCSII) in Beijing second and first epidemics are 326 (on day 24) and 295 (on day 19), respectively. The corresponding blocking rates (Θ_1_) to symptomatic infections reach about 97.48% and 89.32%, respectively.
- The days (denoted by *Day*_100_%) that the blocking rates reached to 100% in Beijing first and second epidemics were on day 98 and on day 28, respectively.
- The days (denoted by *Day*_*clear*_) that all infected individuals were cleared up in Beijing first and second epidemics were on day 140 and on day 56, respectively.
- The numbers of cumulative symptomatic infected individuals in Beijing first and second epidemics are 420 individuals and 335 individuals, respectively,

Although the baseline transmission rate *β*_11_ in the second Beijing epidemic is much higher than the one in the first Beijing epidemic, the higher blocking rate and the recovery rate (after day 27) to the symptomatic infections make that the the second Beijing epidemic ended 84 days earlier than the first epidemic in Beijing and no died infected patients were reported.

## 4 Conclusions

The main contributions of this paper are summarized as follows:

1. It proposes SARDDE models ((1) and (11)) which describe the evolutions of infectious diseases, estimates transmission rates, recovery rates, and blocking rates rate to symptomatic and asymptomatic infections, and symptomatic infected individuals’ death rates.
2. It provides the criterion inequalities for the asymptotical stability of the disease free equilibrium point of SARDDE (see Theorem 1 and Theorem 3).
3. It presents the criterion inequalities for epidemic transmission (see Theorem 2 and Theorem 4) of the symptomatic and asymptomatic infections.
4. It uses two models to simulate the first and second COVID-19 epidemics in Beijing. The simulation results on the end points of transmission intervals were in good agreement with the real word data [17].
5. The simulation results can provide possible interpretations and estimations of the prevention and control measures, and the effectiveness of the treatments to the two epidemics in Beijing.
6. Numerical simulation results suggest that
  i. The transmission rate *β*_11_ of the symptomatic infections caused by the symptomatic individuals in the second Beijing epidemic is much higher than the one in the first Beijing epidemic.
  ii. The blocking rates of 89.32% and 97.48% to the symptomatic infections cannot prevent the spreads of first and second COVID-19 epidemics in Beijing.
  iii. The days that the symptomatic infection blocking rates reached to 100% in Beijing first and second epidemics were on day 98 and on day 28, respectively.
  iv. Using the data form the beginning to the days after about two weeks from the turning points, we can estimate well or approximately the following outcomes of the first or second COVID-19 epidemics in Beijing.
  v. Numerically interpreted that both symptomatic and asymptomatic individuals cause lesser asymptomatic spread than symptomatic spread.
  vi. The lack of the data of the asymptomatic infected individuals in the first COVID-19 in Beijing may prevent the accurate determinations of the number of asymptomatic infected individuals.
  vii. The higher blocking rate and the higher recovery rate (after day 27) to the symptomatic infections make that the the second Beijing epidemic ended 84 days earlier than the first epidemic in Beijing.
  viii. Keeping the blocking rates, recovery rates and death rates reaching the infection turning points would make the numbers of current hospitalized infected individuals reach, on day 140, 208958 individuals and 25017 individuals for the two Beijing epidemics, respectively.
7. Different combinations of the parameters of Equations (1) and (11) may generate similar simulation results. Therefore it needs further study to obtain better parameter combinations to interpret COVID-19 epidemics.
8. The proposed two SARDDEs are simpler than those given in Ref. [11]. However they can better describe and explain the real world data [17].

Because not all infected people can go to the hospital for treatment and be confirmed at the first time. In some cases: adequate resources, no shortage of beds and medical treatment advantages, patients may be left behind when they are discharged from the hospital. Therefore, it does not have very important that the simulation results of the model are required accurately describe every datum reported on the epidemic. Long-term accumulated data, such as the total number of patients and the number of deaths may eliminate short-term deviations. Therefore, the accuracy of predicting long-term epidemics should be the standard for evaluating the rationality of the selected model and unknown model parameters.

When a society cannot withstand high transmission blocking rate load (for example 99%), how can administrative authorities take measures to avoid multiple outbreaks? A recommendation is that the authorities need to at least maintain the prevention and control measures implemented 7 days after reaching the turning point. Administrative authorities need to persuade their society to implement a prevention and control strategy to be withstood until all infection cases are cleared up. It is not a wise strategy to withdraw all prevention and control measures before the number of the all infected people have been cleared up. 100% blocking rate to transmission of COVID-19 infection is the keystone strategy to clear up or reduce spread of an epidemic as early as possible.

The strict prevention and control strategies implemented by Beijing government is not only effective but also necessary. It is expected that the research can provide better understanding, explanation, and dominating the spreads and control measures of epidemics.

## Data Availability

All data cite in this article selected from the web ref. [18] (http://wjw.beijing.gov.cn/

## Funding

The author has not declared a specific grant for this research from any funding agency in the public, commercial or not for profit sectors.

## Conflict of Interest

The author declares no potential conflict of interest.

## Data availability statement

Data are available on reasonable request. Please email the author.

## Ethical Statement

Not applicable/No human participants included.

**Note** The previous versions of the manuscript have been published online [11, 18].

## Footnotes

1 In the cases that some reported data crossed one day, we assign approximately the numbers according to the ratios of time intervals.

## Notes

### Competing Interest Statement

The authors have declared no competing interest.

### Funding Statement

No funding supported in the work.

### Author Declarations

No IRB/oversight body that provided approval or exemption for the research.

### Summary of Updates

Dear The medRxiv team, My previous submitted manuscript is another amended medRxiv paper (ID#: MEDRXIV/2022/2732250). Now the changes in the amended version are given as follows: (1) Three new references are added. (2) A few printed/grammar errors have been corrected.. (3) The styles of sevral references are improved. Lequan

